# Transformer-Based Survival Model for Cardiovascular Risk Prediction from Longitudinal Health Checkup Data

**DOI:** 10.64898/2026.08.24.26361274

**Authors:** Shota Tsurimoto, Akihiro Nomura, Yoshiki Nagata, Masahiro Noguchi, Tadayuki Hirai, Yasuaki Takeji, Hayato Tada, Kenji Sakata, Soichiro Usui, Shogo Okada, Masayuki Takamura

**Author notes:** Corresponding author: Akihiro Nomura, MD, PhD Professor Division of Convergence Science, Kanazawa University Graduate School of Frontier Science Initiative Kakuma-machi, Kanazawa, Ishikawa, 9201192, Japan.

## Abstract

**Background:** Cardiovascular disease (CVD) is a leading global health concern. Traditional models often miss nonlinear dependencies among physiological and behavioral factors. We hypothesized that a Transformer-based deep learning model, which excels at capturing complex patterns in structured data trained on large-scale health check-up records, would improve long-term CVD risk prediction.

**Methods:** We analyzed longitudinal health records (2010–2024) from the Hokuriku Health Service Association (n = 100,056 without baseline CVD; development cohort). Incident CVD was defined as the first self-reported physician diagnosis of heart disease or stroke during the 10-year follow-up and was modeled as right-censored survival data. An external evaluation cohort comprised 79,756 Kanazawa City participants with health records. A Transformer model was trained using anthropometric, laboratory, and self-reported lifestyle data. Benchmarks included Cox regression, XGBoost survival embeddings, multilayer perceptron, the Framingham Risk Score, and the Hisayama Risk Score. Performance was evaluated using time-dependent area under the receiver operating characteristic curve (ROC-AUC) with a primary focus on the 10-year ROC-AUC, precision–recall AUC (PR-AUC), and concordance index (C-index). Interpretability was assessed through SHapley Additive exPlanations (SHAP) and a Feature-level Attention Network (FAN), visualizing the top 12 SHAP-ranked features to highlight key interactions.

**Results:** In the development cohort, 4,113 CVD events (4.1%) occurred. The Transformer model achieved the best internal performance: 10-year ROC-AUC 0.821 (95% confidence interval [CI], 0.816– 0.826), PR-AUC 0.427 (CI, 0.419–0.435), and C-index 0.781 (CI, 0.775–0.787). Performance remained robust externally (21,179 CVD events, 26.6%): ROC-AUC, 0.762; PR-AUC, 0.500; and C-index, 0.744. Regarding interpretability, SHAP identified age, electrocardiogram abnormality, antihypertensive medication, and sex as the most critical predictors. Notably, FAN elucidated the prognostic value of self-reported lifestyle factors. For example, daily exercise and weight gain modulated the model’s assessment of age-related risk. Within the attention network, age served as a central hub, linking these behavioral habits with physiological features.

**Conclusion:** The Transformer-based model outperformed conventional methods in predicting long-term CVD risk. Model interpretation demonstrated the predictive utility of self-reported lifestyle factors, such as weight gain and daily exercise. These findings may support personalized CVD prevention and population-level risk stratification using routinely collected health checkup data.

**Clinical Perspective:** *What Is New?:*

- A Transformer-based survival model outperformed conventional models and clinical risk scores for 10-year cardiovascular disease risk prediction in both internal and external evaluations.
- Model interpretation showed that the model used both established clinical risk factors and lifestyle factors, including daily exercise and weight gain.

*What Are the Clinical Implications?:*

- Routinely collected health checkup data may support long-term cardiovascular disease risk stratification.
- This model may help identify high-risk individuals and support targeted lifestyle guidance and preventive care.

## Introduction

Cardiovascular disease (CVD) remains one of the leading causes of death worldwide, accounting for more than 17 million deaths each year.^1^ Because incident CVD places a major burden on individuals and society, identifying high-risk individuals early and providing timely preventive interventions are essential for CVD prevention.^2^

Traditionally, high-risk individuals have been identified using risk scores such as the Framingham Risk Score (FRS) and the Hisayama Risk Score (HRS).^3,4^ These scores estimate long-term CVD risk based on a limited set of conventional predictors, including age, sex, blood pressure, lipid levels, smoking, and diabetes. Although these models are simple and highly interpretable, their predictive performance can be somewhat limited.^5–7^ This limitation is largely due to the use of Cox proportional hazards models, which generally assume proportional hazards over time and relatively simple covariate effects. As a result, these scores may not fully capture nonlinear relationships, complex interactions, or changes in risk effects over time.^8,9^ At the same time, growing evidence suggests that modifiable health behaviors, including alcohol consumption, physical inactivity, insufficient sleep, and unhealthy diet, are associated with CVD outcomes.^10–12^ These findings highlight the need for advanced risk models that can integrate a wide range of clinical, laboratory, and lifestyle factors.

Machine learning (ML) approaches offer a promising solution to address these limitations. In particular, deep learning models can automatically learn complex nonlinear and higher-order relationships across a diverse set of input variables.^13,14^ However, when applied to conventional tabular data, deep learning models have often failed to consistently outperform tree-based methods or traditional statistical models, frequently yielding only comparable performance.^15^

To overcome this challenge, we focused on Transformer-based neural networks, originally developed for natural language processing tasks.^16^ By treating each input feature as a token, Transformer models can learn contextual dependencies among variables through self-attention mechanisms. Several recent studies have reported strong performance of Transformers on tabular prediction tasks.^17,18^ Therefore, we hypothesized that a Transformer-based model would be effective for predicting long-term CVD risk from structured health check-up data.

In this study, we aimed to develop a Transformer-based survival prediction model for long-term CVD risk and compare its performance against conventional risk prediction models. The proposed model integrated standard clinical and laboratory parameters alongside self-reported lifestyle factors, including physical activity, sleep, and dietary habits. The model was initially developed using a large-scale cohort of over 1.2 million records with more than 10 years of longitudinal health checkup records provided by the Hokuriku Health Service Association. We subsequently validated its performance using an independent, external evaluation cohort of approximately 800,000 records with 15-year longitudinal records from the Kanazawa City Medical Association.

## Methods

### Study overview

We developed and evaluated a Transformer-based model for predicting 10-year CVD risk using two independent, longitudinal real-world health checkup datasets from the Japanese general population. These datasets included clinical parameters, laboratory measurements, and lifestyle factors. The performance of the proposed model was compared with established statistical and machine learning methods, as well as conventional CVD risk scores as the FRS and the HRS. To enhance transparency and clinical applicability, we further applied interpretability methods to identify key contributors to the predicted risk.

The study was conducted in accordance with the Declaration of Helsinki and the Ethical Guidelines for Medical and Biological Research Involving Human Subjects in Japan. Because of the retrospective design and the use of deidentified, anonymized secondary data, the requirement for informed consent was waived. The study protocol was approved by the Medical Ethics Committee of Kanazawa University (Research Protocol Number: 2019-080 [113122]).

### Study population and data sources

The study flowchart is summarized in **Figure 1**. The development cohort consisted of 1,220,950 health checkup records from 294,698 individuals, derived from the database of employer-mandated annual health examinations conducted by the Hokuriku Health Service Association (Toyama, Japan) between April 2010 and March 2024 (HHSA cohort). These health checkups were performed at affiliated hospitals or clinics and included standardized blood and urine tests, anthropometric measurements, and a structured self-administered lifestyle questionnaire. For model development, participants in the HHSA cohort were randomly split into a training/validation subset (80%) and an internal test subset (20%).

**Figure 1.**
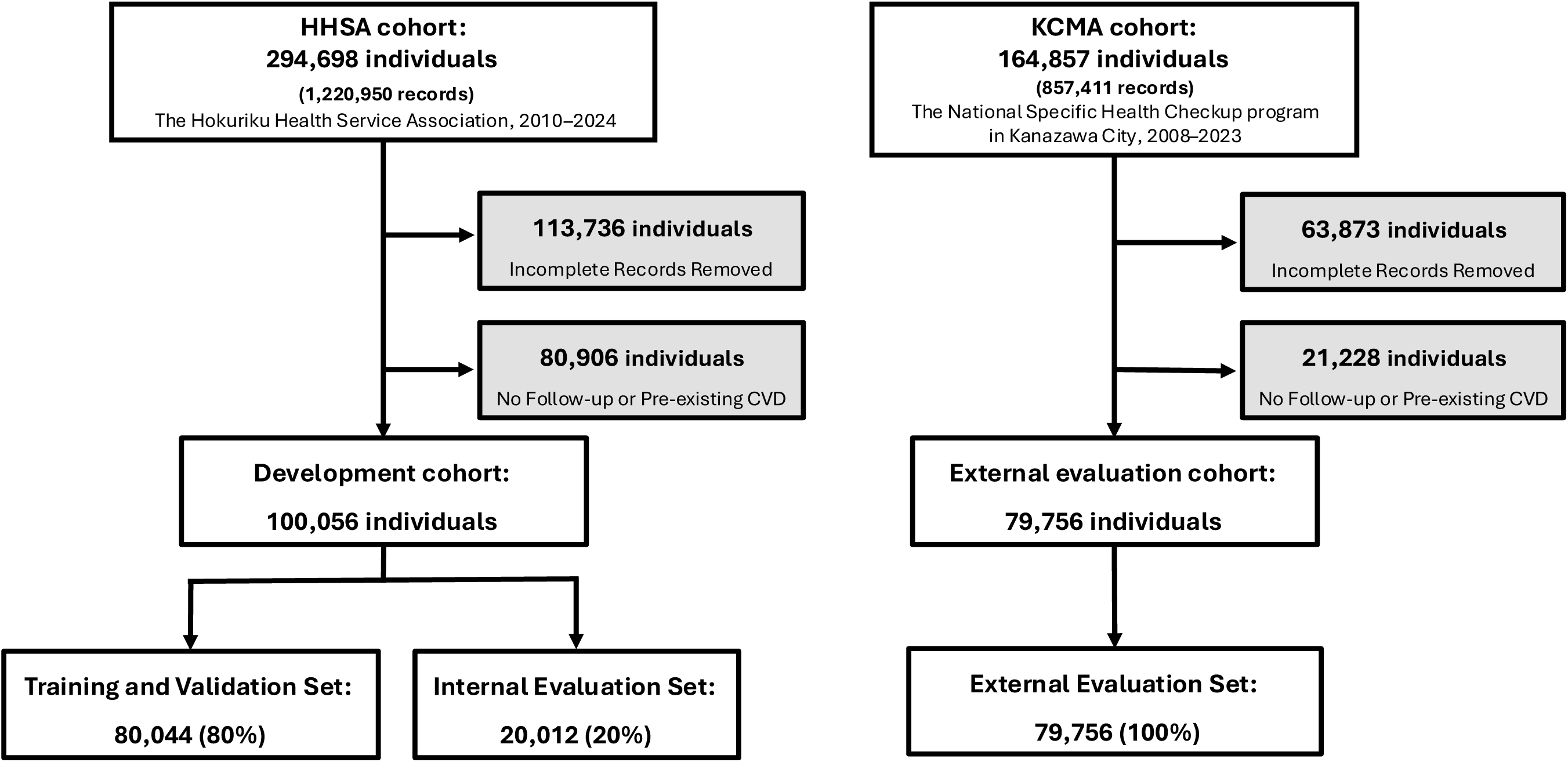
Study flowchart. Participant flow diagram for the development and external evaluation cohorts. The development cohort comprised individuals undergoing annual health examinations conducted by the Hokuriku Health Service Association (HHSA), and the external evaluation cohort included participants from the Kanazawa City Medical Association (KCMA).

To further assess external validity, we used an independent external evaluation cohort comprising 857,411 health checkup records from 164,857 participants aged ≥40 years in Kanazawa City. These data were obtained from the national Specific Health Checkup program conducted by the Kanazawa City Medical Association (Ishikawa, Japan) between April 2008 and March 2023 (KCMA cohort). This program primarily targets individuals aged 40–74 years enrolled in the National Health Insurance system, those aged ≥75 years enrolled in the Late-Stage Elderly Healthcare System, and individuals receiving public assistance. The clinical examination protocol and self-reported lifestyle assessments in this program were largely harmonized with those used in the HHSA cohort.

### Definitions and follow-up

The primary outcome were the occurrence of CVD events. Events were ascertained by longitudinally tracking responses to an annual self-administered questionnaire item included in each health checkup: “Have you ever been told by a physician that you have heart disease or stroke?” A CVD event was defined as the first transition from a “no” response at baseline to a “yes” response at any subsequent visit during follow-up. Self-reported CVD events encompassed myocardial infarction, angina, heart failure, ischemic stroke, and hemorrhagic stroke. The event year was defined as the calendar year in which CVD was first reported.

Baseline was defined as the date of the first health checkup during the study period for each participant. The full definitions of each baseline characteristic are provided in **Supplementary Table S1**. Follow-up time was calculated from baseline to the earliest of the following: 1) the year of CVD occurrence; 2) the date of the last health checkup; or 3) 10 years after baseline. Participants who did not report a CVD event during follow-up were treated as censored at their last available health checkup. These definitions were applied consistently across both cohorts.

#### Inclusion and exclusion criteria

We included participants if they had at least one baseline health checkup during the study period, all baseline covariates required for model construction, no self-reported history of CVD at baseline, and follow-up information sufficient to determine either incident CVD or censoring status. Conversely, we excluded individuals if they had missing values for any required baseline variable, missing outcome data, no follow-up assessment after baseline, or a baseline history of heart disease or stroke. These inclusion and exclusion criteria were prespecified and applied consistently to both the HHSA development cohort and the KCMA external evaluation cohort prior to model development and evaluation.

### Feature set and preprocessing

The prediction model was trained using 43 features extracted from routine health checkup records (**Supplementary Table S1**). These features comprised demographic variables, anthropometric measurements, vital signs, clinical laboratory biomarkers, complete blood count, urinalysis findings, and self-reported medication use (antihypertensive, glucose-lowering, or lipid-lowering agents). Electrocardiogram abnormalities were interpreted by cardiologists based on the Minnesota Code (MC) and were categorized into three levels according to the severity of abnormalities (normal, borderline, and abnormal) and incorporated as a categorical variable. For instance, mild ST-segment depression (MC 4-6), complete right bundle branch block (MC 7-2), and left axis deviation (MC 2-1) were classified as borderline abnormal, whereas marked ST-segment depression (MC 4-1), abnormal Q waves (MC 1-2), and frequent premature atrial contractions (MC 8-1-1) were classified as abnormal. Additionally, detailed lifestyle factors derived from structured questionnaire were included, such as smoking status, alcohol intake, physical activity, sleep quality, excess weight gain since age of 20, dietary habits, and motivation to improve lifestyle behaviors.

### Model development and benchmarks

We developed a deep learning survival model based on a Transformer architecture designed to capture complex dependencies among tabular features via multi-head self-attention (**Figure 2**). Continuous and categorical features were first mapped into dense vector representations using numerical and categorical embedding layers, respectively. These embedded feature tokens were then processed by the Transformer architecture consisting of three stacked Transformer encoder blocks, each with four attention heads of 64 dimensions per head. The output of the Transformer backbone was passed through two fully connected layers (128 and 64 units, respectively) to generate the final risk predictions. Residual connections and layer normalization were incorporated throughout the network to ensure stable training. The model was trained to predict discrete-time hazard functions over a 10-year horizon using a negative log-likelihood loss function adapted for right-censored survival data, as proposed by Gensheimer and Narasimhan.^19^

**Figure 2.**
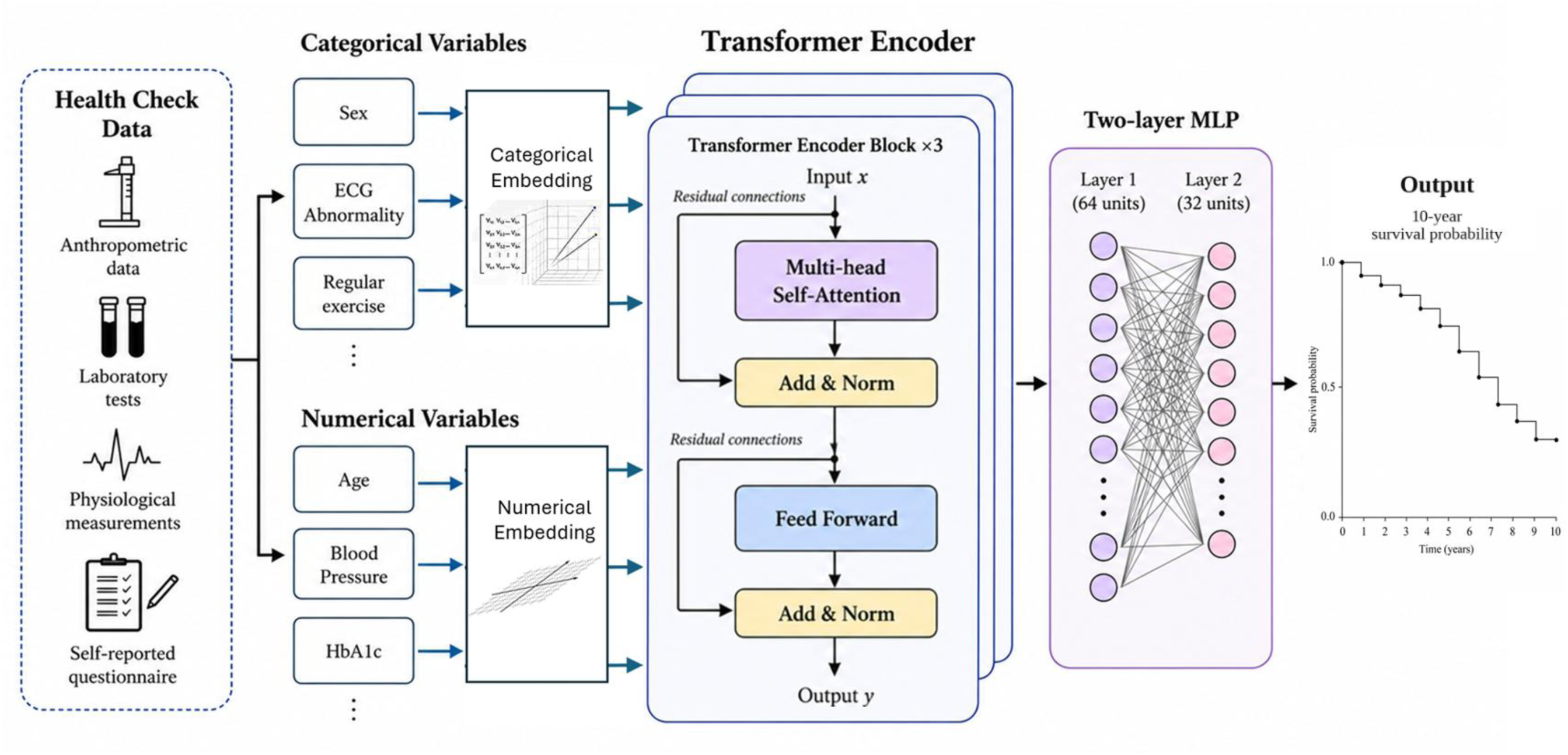
Overview of the Transformer-based survival prediction model for CVD. Categorical and numerical variables derived from health check data are embedded separately and represented as feature tokens. These tokens are processed by a Transformer encoder to model complex dependencies among variables through multi-head self-attention. The encoded representations are then combined and passed through a multilayer perceptron, yielding estimated survival probabilities over time, including the predicted 10-year survival probability.

For model development, we used the training/validation subset (80% of participants) from the development cohort. Hyperparameters were tuned using stratified 5-fold cross-validation to mitigate overfitting. We used the Adam optimizer with a learning rate of 0.001 and a weight decay of 1×10⁻⁵. Models were trained with a batch size of 256, and early stopping based on validation loss was applied (patience = 10 epochs).^20^ Dropout rates and model depth were optimized via random search. All models were implemented in PyTorch (v1.13) with a fixed random seed (seed = 0) to ensure reproducibility.

For evaluation, we implemented several comparator models using the same feature set and evaluation protocol: I) a three-layer multilayer perceptron (MLP) as a standard deep learning baseline without attention; II) a survival-adapted XGBoost model trained using the Cox partial likelihood loss (XGBSE)^21^; a Cox proportional hazards model ^22^; IV) the FRS, a widely used tool for estimating 10-year CVD risk^3^; and V) the HRS, a 10-year CVD risk prediction model developed specifically for Japanese adults.^4^

### Statistical analysis

For model input, all continuous variables were standardized using z-scores based on the mean and standard deviation calculated from the training data. Categorical variables were label-encoded into integer values according to predefined categories. Model performance was primarily compared using the area under the receiver operating characteristic curve (ROC–AUC) at 10 years^23^, consistent with the primary evaluation metrics of the Framingham and Hisayama risk scores. To assess performance over the entire follow-up period, we additionally calculated Harrell’s concordance index (C-index)^24^ and time-dependent ROC–AUC values for each year within the 10-year follow-up.^25^ Given the relatively low incidence of CVD events, we also evaluated the time-dependent area under the precision–recall curve (PR–AUC)^26^ to provide a more informative assessment of positive predictive performance under class imbalance. Model calibration was assessed by comparing the Brier score across models.^27^ For all performance metrics, 95% confidence intervals (CIs) were estimated using 1,000 bootstrap resamples.^28^ Statistical analyses were conducted in Python using the lifelines (v0.27) and scikit-survival (v0.17.2) libraries for survival analysis.

### Model Interpretation

We employed two complementary approaches to evaluate the interpretability of the proposed Transformer-based prediction model. First, we applied an extension of the Kernel SHAP algorithm^29^ to survival outcomes to estimate the marginal contribution of each input feature to the model predictions. Second, to better understand the representations learned in the Transformer attention blocks, we proposed a Feature-level Attention Network (FAN). This network visualizes information flow among features as a directed graph, where nodes represent features and edges represent attention-based interactions. While this approach built upon prior work regarding attention visualization in Transformer models^30–32^, we reformulated the attention patterns specifically as a feature-level network. In the FAN approach, we examined how features with high SHAP contributions attended to one another using the query-side attention weights associated with each feature. To reduce noise, we retained only the top 10% of attention queries as edges. We then computed betweenness centrality on the resulting network to identify the top five critical nodes. Betweenness centrality was defined as the proportion of shortest paths between all pairs of nodes that pass through a given node, effectively reflecting its importance as an information hub within the network.^33^ Network construction and visualization were implemented in Python using the networkx library (v3.2).

## Results

In the HHSA cohort, we initially identified 1,220,950 health checkup records from 294,698 individuals. Individuals were linked longitudinally using unique participant identifiers. We then applied the prespecified exclusion criteria: 113,736 participants were excluded due to missing variables in one or more required variables, and an additional 80,906 participants were excluded because they lacked follow-up assessments after baseline or had a history of CVD at baseline. The resulting HHSA analytic cohort comprised 100,056 participants. Similarly, in the KCMA cohort, we obtained 857,411 health checkup records from 164,857 participants. Applying identical exclusion criteria, 63,873 participants were excluded due to missing values in required variables, and a further 21,228 participants were excluded due to a lack of follow-up assessments or a baseline history of CVD. The resulting KCMA analytic cohort comprised 79,756 participants (**Figure 1**).

For model development, we used the HHSA cohort. We randomly partitioned the HHSA participants into a training/validation set (80%) for model training and hyperparameter tuning, and a holdout testing set (20%) for internal evaluation. The final trained models were subsequently applied to the independent KCMA cohort for external evaluation.

### Baseline characteristics

**Table 1** summarizes the major baseline characteristics of the HHSA development cohort and the KCMA external evaluation cohort. Full feature demographics are provided in **Supplementary Table 2**. Participants in the HHSA cohort were generally younger and more frequently male compared to the KCMA cohort. The KCMA cohort exhibited a less favorable cardiometabolic risk profile, characterized by higher mean systolic and diastolic blood pressure, triglycerides, plasma glucose, and hemoglobin A1c (HbA1c) levels, and lower high-density lipoprotein cholesterol, and correspondingly higher proportions of alongside lower high-density lipoprotein cholesterol levels. Consistent with this higher-risk profile, the antihypertensive and lipid-lowering medication use. Despite a shorter mean follow-up, the KCMA cohort had a higher cumulative incidence of CVD.

**Table 1.** Key baseline characteristics of HHSA and KCMA cohorts.

|  | HHSA dataset (n=100,056) | KCMA dataset (n=79,756) |
| --- | --- | --- |
| **Age, years** | 45 ± 11 | 68 ± 10 |
| **Male sex, n (%)** | 60,957 (60.9%) | 31,260 (39.2%) |
| **Medication, n (%)** |  |  |
| Antihypertensive agents | 9,049 (9.0%) | 31,953 (40.1%) |
| Insulin use | 2,982 (3.0%) | 6,827 (8.6%) |
| Lipid-lowering agents | 3,667 (3.7%) | 19,772 (24.8%) |
| **Blood test, mean ± SD** |  |  |
| Total cholesterol, mg/dL | 206 ± 38 | 205 ± 35 |
| LDL-C, mg/dL | 121 ± 32 | 121 ± 31 |
| Glucose, mg/dL | 96 ± 23 | 103 ± 30 |
| HbA1c, % | 5.6 ± 0.7 | 5.5 ± 0.7 |
| Creatinine, mg/dL | 0.8 ± 0.2 | 0.7 ± 0.2 |
| eGFR, ml/min/1.73m<sup>2</sup> | 82 ± 15 | 72 ± 16 |
| **Electrocardiogram abnormality, n (%)** |  |  |
| Normal | 79,629 (79.6%) | 57,773 (72.4%) |
| Borderline | 12,853 (12.8%) | 14,419 (18.1%) |
| Abnormal | 7,574 (7.5%) | 7,564 (9.5%) |
| **Self-reported lifestyle, n (%)** |  |  |
| Smoking, n (%) | 29,491 (29.5%) | 10,423 (13.1%) |
| Weight gain since age of 20 | 29,242 (29.2%) | 24,161 (30.3%) |
| Daily Exercise | 21,002 (21.0%) | 42,055 (52.7%) |
| Eating Speed |  |  |
| Slow | 34,366 (34.3%) | 23,117 (29.0%) |
| Normal | 57,260 (57.2%) | 45,596 (57.2%) |
| Fast | 8,430 (8.4%) | 11,043 (13.8%) |
| **CVD events, n (%)** | 4,113 (4.1%) | 21,179 (26.6%) |
| **Mean follow-up duration, years** | 4.9 ± 7.1 | 7.1 ± 8.0 |

**Abbreviations:** ECG, electrocardiogram; eGFR, estimated glomerular filtration rate; HbA1c, glycated hemoglobin A1c; HHSA, Hokuriku Health Service Association; KCMA, Kanazawa City Medical Association; LDL-C, low-density lipoprotein cholesterol; SD, standard deviation.

### Model Performance

First, we evaluated the 10-year CVD prediction performance of each model using the internal testing subset of the HHSA development cohort, comprising 20% of the HHSA participants. In this testing subset, the Transformer-based model achieved the highest 10-year ROC–AUC of 0.821 (95% CI, 0.816– 0.826), outperforming SimpleMLP (0.788), XGBoost (0.794), the Cox model (0.756), the FRS (0.719), and the HRS (0.724) (**Figure 3A**). The Transformer-based model also demonstrated the highest 10-year PR–AUC of 0.427 (95% CI, 0.419–0.435), surpassing XGBoost (0.400), the Cox model (0.381), SimpleMLP (0.338), the HRS (0.304), and the FRS (0.293). Similarly, the Transformer-based model yielded the highest C-index of 0.781 (95% CI, 0.775–0.787), followed by XGBoost (0.770), the Cox model (0.761), SimpleMLP (0.730), the HRS (0.729), and the FRS (0.727). Regarding calibration, the Transformer-based model showed a Brier score of 0.031 (95% CI, 0.030–0.032), which was comparable to those of SimpleMLP, the Cox model, and the HRS, and lower than those of XGBoost (0.049) and the FRS (0.037) (**Table 2**).

**Figure 3.**
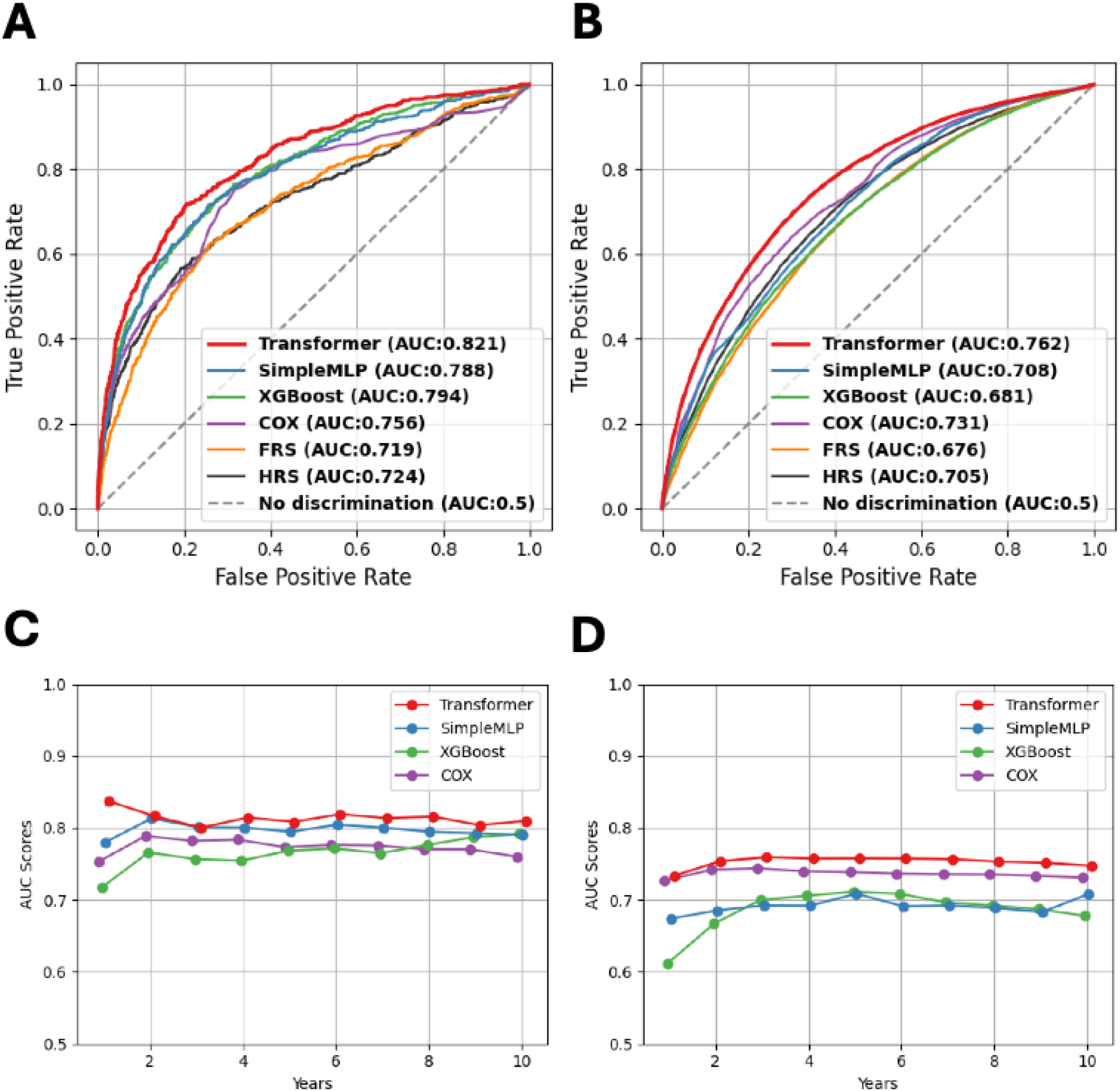
Performance comparison of the Transformer model and comparator models in the internal testing subset and the external evaluation cohort. (**A, B**) ROC curves for the Transformer model, SimpleMLP, XGBoost, Cox model, FRS, and HRS in the HHSA internal testing subset (**A**) and the KCMA external evaluation cohort (**B**). (**C, D**) Time-dependent ROC–AUCs from 1 to 10 years for the Transformer model, SimpleMLP, XGBoost, and Cox model in the HHSA internal testing subset (**C**) and the KCMA external evaluation cohort (**D**).

**Table 2.** Model performance metrics in the HHSA testing subset and the KCMA external evaluation cohort.

| Model | 10-year ROC-AUC<br>[95% CI] | 10-year PR-AUC<br>[95% CI] | C-index<br>[95% CI] | Brier score<br>[95% CI] |
| --- | --- | --- | --- | --- |
| **HHSA internal testing subset** |  |  |  |  |
| Transformer model | 0.821[0.816-0.826] | 0.427[0.419-0.435] | 0.781[0.775-0.787] | 0.031[0.030-0.032] |
| SimpleMLP model | 0.788[0.782-0.795] | 0.338[0.328-0.347] | 0.730[0.722-0.737] | 0.031[0.030-0.032] |
| XGBoost model | 0.794[0.790-0.797] | 0.400[0.388-0.411] | 0.770[0.761-0.778] | 0.049[0.047-0.050] |
| Cox model | 0.756[0.753-0.759] | 0.381[0.373-0.389] | 0.761[0.757-0.765] | 0.031[0.031-0.032] |
| Framingham Risk Score | 0.719[0.711-0.758] | 0.293[0.272-0.313] | 0.727[0.700-0.754] | 0.037[0.035-0.039] |
| Hisayama Risk Score | 0.724[0.704-0.759] | 0.304[0.282-0.326] | 0.729[0.697-0.760] | 0.031[0.029-0.033] |
| **KCMA external evaluation cohort** |  |  |  |  |
| Transformer model | 0.762[0.761-0.763] | 0.500[0.498-0.502] | 0.744[0.744-0.745] | 0.168[0.167-0.168] |
| SimpleMLP model | 0.708[0.707-0.709] | 0.326[0.324-0.327] | 0.657[0.656-0.658] | 0.172[0.171-0.172] |
| XGBoost model | 0.681[0.680-0.682] | 0.417[0.414-0.419] | 0.704[0.703-0.705] | 0.191[0.191-0.192] |
| Cox model | 0.731[0.731-0.732] | 0.439[0.437-0.441] | 0.714[0.713-0.714] | 0.149[0.149-0.150] |
| Framingham Risk Score | 0.676[0.665-0.680] | 0.321[0.316-0.325] | 0.655[0.651-0.658] | 0.149[0.147-0.151] |
| Hisayama Risk Score | 0.705[0.694-0.711] | 0.340[0.336-0.343] | 0.683[0.680-0.685] | 0.149[0.147-0.151] |

**Abbreviations:** AUC, area under the curve; CI, confidence interval; C-index, Harrell's concordance index; FRS, Framingham Risk Score; HHSA, Hokuriku Health Service Association; HRS, Hisayama Risk Score; KCMA, Kanazawa City Medical Association; MLP, multilayer perceptron; PR-AUC, precision-recall area under the curve; ROC, receiver operating characteristic; XGBoost, extreme gradient boosting.

Next, we evaluated the models on the independent KCMA external evaluation cohort. The Transformer-based model again achieved the highest 10-year ROC–AUC of 0.762 (95% CI, 0.761–0.763), outperforming the Cox model (0.731), SimpleMLP (0.708), the HRS (0.705), XGBoost (0.681), and the FRS (0.676) (**Figure 3B**). Furthermore, the Transformer-based model attained the highest 10-year PR– AUC of 0.500 (95% CI, 0.498–0.502), and a C-index of 0.744 (95% CI, 0.744–0.745), consistently exceeding all baseline models. Conversely, the Brier score of the Transformer-based model in this cohort was 0.168 (95% CI, 0.167–0.168), which was higher than those of the Cox model, the FRS, and the HRS (each 0.149), but lower than those of SimpleMLP (0.172) and XGBoost (0.191) (**Table 2**).

Across both the development and external evaluation cohorts, the Transformer-based model maintained robust discrimination across prediction horizons ranging from 1 to 10 years (**Figure 3C and 3D**).

### Model interpretability

Finally, we assessed the interpretability using SHAP and FAN. **Figure 4A** illustrates the top 12 predictors ranked by their mean absolute SHAP values. Age was the most influential feature, followed by ECG abnormalities, antihypertensive medication use, sex, and glycemic indices. Notably, modifiable lifestyle factors, such as daily physical activity, excess weight gain since age of 20, and fast eating speed demonstrated substantial contributions to CVD risk prediction. These were comparable to established clinical and physiological markers as total cholesterol, low-density lipoprotein cholesterol, HbA1c, or estimated glomerular filtration rate. Furthermore, FAN visualization revealed that age emerged as the primary node receiving the strongest attention from multiple other features, while ECG abnormalities, HbA1c, antihypertensive treatment history, and habitual physical activity occupied central hub positions within the network (**Figure 4B**).

**Figure 4.**
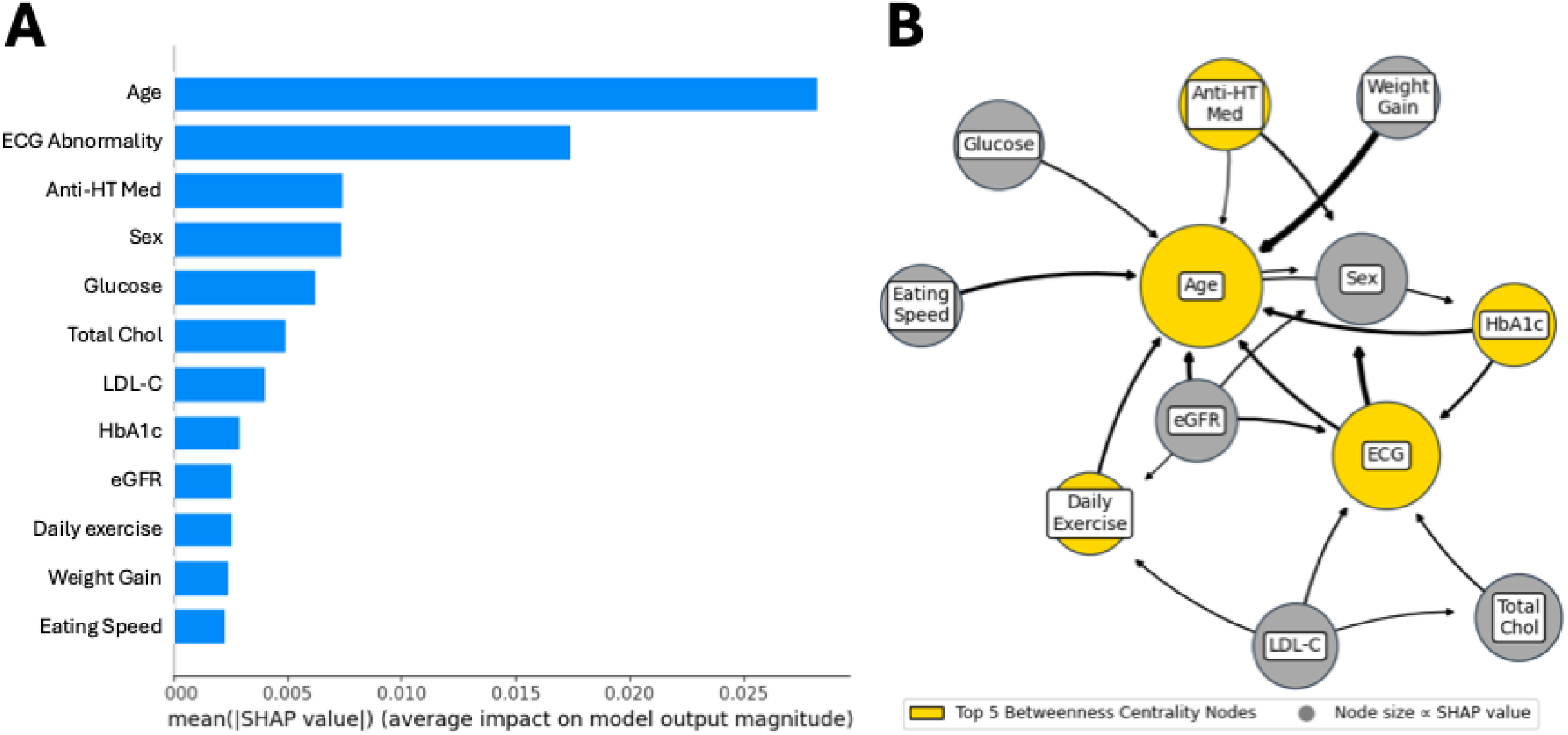
Model interpretability analysis using SHAP values and the Feature-level Attention Network. **(A)** Ranking of the top 12 features according to mean absolute SHAP values, indicating their average contribution to the model output. Age showed the greatest contribution, followed by ECG abnormalities and current use of antihypertensive medication. (**B**) Feature-level Attention Network illustrating how the top 12 SHAP-ranked features interact within the model. Node size is proportional to the SHAP value of each feature, reflecting its overall importance. Edge thickness represents the strength of the attention-based interaction between features. Yellow nodes indicate the top five features ranked by betweenness centrality, highlighting variables that serve as central hubs for information flow within the network.

## Discussion

In this study, we developed a Transformer-based survival prediction model for CVD using two large, longitudinal, population-based cohorts in Japan and evaluated its predictive performance. The Transformer-based model consistently demonstrated strong performance in both internal and external evaluations, outperforming conventional approaches such as the Cox proportional hazards model, tree-based methods, standard neural networks, and widely used CVD risk scores as the FRS or HRS. Furthermore, our model successfully integrated lifestyle factors such as habitual physical activity, which have generally been overlooked by conventional CVD risk prediction models, thereby improving predictive performance.

Our findings provide several key insights. First, the proposed Transformer-based neural network model consistently outperformed conventional prediction methods. This finding aligns with previously reported results on Transformer-based models for tabular data.^17,34,35^ For example, Tab-Transformer improved the average ROC–AUC by at least 1.0% compared to conventional deep learning methods across 15 publicly available datasets, and showed a ROC–AUC increase of 2.1% when combined with semi-supervised pre-training.^17^ Similarly, Transformer-based models, such as FT-Transformer (Feature Tokenizer Transformer) and SAINT (the Self-Attention and Intrasample Attention Transformer), have been shown to consistently improve upon existing deep learning methods.^34,35^ The superior performance of our Transformer-based survival model suggests that self-attention may help capture complex interactions among features in tabular health checkup data, thereby improving long-term CVD risk prediction.

Second, model interpretation using SHAP and FAN suggested that our model effectively incorporated lifestyle factors, including habitual physical activity, which have not been fully integrated into conventional CVD risk prediction models. In the SHAP analysis, age, electrocardiographic abnormalities, blood pressure–related factors, lipid markers, renal function, and glycemic indices emerged as influential predictors. These findings were broadly consistent with the key predictors identified in the Hisayama and Suita CVD risk models, suggesting that the model appropriately learned established clinical determinants of CVD risk.^4,36^ In addition to these conventional factors, our analysis showed that lifestyle-related variables, such as exercise habits, weight gain from the age of 20 years, and eating speed, also made substantial contributions to prediction. Notably, FAN showed that exercise habits acted as a hub connecting age with risk-related factors such as renal function and cholesterol. This suggests that physical activity may contribute to CVD risk not only as an independent factor, but also through its relationships with other clinical and metabolic risk factors. Previous machine learning studies using health checkup data have also emphasized the prognostic value of lifestyle factors.^37–39^ In addition, epidemiological studies have consistently shown that higher levels of physical activity are associated with lower risks of both incident CVD and CVD mortality.^40^ Taken together, these findings suggest that our model not only reproduced established risk factors, but also captured more complex CVD risk structures shaped by interactions between physiological markers and lifestyle behaviors.

Third, this study showed that annual health checkup data can provide a practical basis for long-term CVD risk prediction. Compared with electronic medical records from hospitals and clinics, health checkup data collected through national health programs have not traditionally been regarded as a major research resource. However, these data have several important strengths, including standardized measurement protocols, broad coverage of community residents and workers, and the feasibility of long-term follow-up. Previous studies support these advantages. For example, in a large Japanese occupational cohort of 155,108 individuals, a random survival forest model using annual health checkup data achieved favorable performance in predicting 6-year CVD events.^41^ Similarly, in a nationwide health checkup cohort in South Korea, a deep learning model using repeated health checkup data demonstrated good discrimination for CVD risk prediction.^42^ These findings support the usefulness of health checkup data for population-level CVD risk stratification. Our study further reinforces the view that health checkup databases are not merely screening records, but valuable large-scale resources for long-term prognostic modeling. Future research should focus on expanding and leveraging such datasets to implement predictive models in preventive medicine.

Several limitations should be noted. First, CVD outcomes were based on self-reported physician diagnoses, meaning outcome misclassification cannot be entirely ruled out. However, previous studies in Japan have reported acceptable agreement between self-reported stroke or myocardial infarction and registry-based diagnoses, supporting the validity of self-reported outcomes in epidemiological research.^43^ Second, although our Transformer model exhibited superior discriminative performance compared to the comparator models, its calibration was less favorable in the external evaluation cohort. This discrepancy is partly attributable to differences in baseline characteristics between the development and external evaluation cohorts, which may have induced calibration drift despite preserved discrimination. Third, because both cohorts were drawn from geographically adjacent areas in Japan, our findings may not be fully generalizable to other populations, regions, or healthcare settings. Although major overlap between the cohorts is unlikely, minor unintentional participant overlap over more than 10 years of follow-up cannot be completely excluded. Fourth, SHAP and FAN are post-hoc explainability tools that describe how trained predictive models behave. Therefore, the highlighted features should not be directly interpreted as causal CVD risk factors. Recent studies in explainable AI have shown that Shapley value– based methods explain approximate model behavior rather than true causal effects, and that their interpretation is limited when features are strongly correlated.^44^ In addition, attention weights do not necessarily indicate the true contribution of features to model predictions.^45,46^ Therefore, attention-based explanations should be interpreted with caution. Our visualization results should be viewed as hypothesis-generating findings that suggest possible feature-interaction patterns used by the model. Further external validation and causal inference studies are needed before these findings can be translated into clinical decision-making.

## Conclusion

We developed a Transformer-based survival prediction model for CVD that consistently outperformed conventional models and widely used clinical risk scores. Interpretability analyses using SHAP and FAN showed that the model captured established clinical risk factors as well as lifestyle-related information, particularly habitual physical activity. These findings support the utility of Transformer-based deep survival modeling for CVD risk stratification using routinely collected longitudinal health checkup data and may help inform personalized lifestyle interventions and proactive CVD prevention.

## Data Availability

The data are not publicly available because they contain potentially identifiable health information and are subject to data-use agreements with the data providers. Access to the data requires approval from the respective data providers and relevant ethics committees.

## Acknowledgments

We thank all participants from the Hokuriku Health Service Association and the municipal health departments of Toyama Prefecture and Kanazawa City for their assistance in data collection and management. We are also grateful to Yoshitaka Sakikawa at the Kanazawa Medical Association for his technical support, and Yuki Kosaka at NEC Corporation for his valuable advice. During the preparation of this manuscript, Gemini (Google LLC, CA, the USA) was used to support English language editing. We reviewed the edited text after using the tool and took full responsibility for the final content of the manuscript.

## Data Sharing

The code for this article is hosted at GitHub: https://github.com/tsurimoto828/cvd-transformer-survival The datasets analyzed in the current study are not publicly available due to data use agreements with local health authorities and privacy protection regulations. Requests for access to anonymized, aggregated data may be directed to the corresponding author and will be considered on a case-by-case basis in accordance with institutional and ethical guidelines.

---

**Supplementary Table 1.** Definition of each baseline characteristic obtained from HHSA and KCMA cohorts.

| Features | Type | Definition |
| --- | --- | --- |
| Age | Continuous | Age (years) |
| Height | Continuous | Body height (cm) |
| Weight | Continuous | Body weight (kg) |
| BMI | Continuous | Body mass index ($\text{kg}/\text{m}^2$) |
| Waist | Continuous | Waist circumference (cm) |
| SBP | Continuous | Systolic blood pressure (mmHg) |
| DBP | Continuous | Diastolic blood pressure (mmHg) |
| RBC | Continuous | Red blood cell count ($\times 10^6/\mu\text{L}$) |
| Hemoglobin | Continuous | Hemoglobin concentration (g/dL) |
| Hematocrit | Continuous | Hematocrit (%) |
| AST | Continuous | Aspartate aminotransferase (U/L) |
| ALT | Continuous | Alanine aminotransferase (U/L) |
| GGT | Continuous | Gamma-glutamyl transferase (U/L) |
| Total Chol | Continuous | Total cholesterol (mg/dL) |
| HDL-C | Continuous | High-density lipoprotein cholesterol (mg/dL) |
| Triglyceride | Continuous | Triglycerides (mg/dL) |
| LDL-C | Continuous | Low-density lipoprotein cholesterol (mg/dL) |
| Glucose | Continuous | Fasting plasma glucose (mg/dL) |
| HbA1c | Continuous | Glycated hemoglobin A1c (%) |
| Creatinine | Continuous | Serum creatinine (mg/dL) |
| eGFR | Continuous | Estimated glomerular filtration rate (mL/min/1.73 m<sup>2</sup>) |
| Sex | Categorical | Sex |
| Urine Protein | Categorical | Protein in urine on dipstick urinalysis |
| Urine Glucose | Categorical | Glucose in urine on dipstick urinalysis |
| ECG abnormality | Categorical | Abnormal findings on a resting 12-lead ECG interpreted by a cardiologist |
| Fundus of eye test | Categorical | Presence of abnormal findings on fundus examination at the health checkup |
| Anti-HT Med | Categorical | Current use of antihypertensive medication |
| Insulin Med | Categorical | Current use of insulin therapy for diabetes |
| Anti-Chol Med | Categorical | Current use of lipid-lowering medication |
| Past Anemia | Categorical | History of anemia |
| Smoker | Categorical | Current cigarette smoking |
| Weight gain since age of 20 | Categorical | Weight gain of more than 10 kg in body weight since the age of 20 |
| Regular Exercise | Categorical | Habit of exercising for at least 30 minutes at a time, at least twice a week, continued for at least 1 year |
| Daily Exercise | Categorical | Habit of walking or performing equivalent physical activity for at least 1 hour every day |
| Fast Walking | Categorical | Usual walking speed faster than that of people of the same sex and age |
| Eating Speed | Categorical | Usual speed of eating meals, categorized as slow, medium, or fast |
| Late Meals | Categorical | Habit of eating dinner within 2 hours of going to bed at least three times per week |
| Skip Breakfast | Categorical | Habit of skipping breakfast at least three times per week |
| Habitual Drinker | Categorical | Habit of drinking alcoholic beverages almost every day |
| Daily alcohol intake | Categorical | Average amount of alcohol consumed on a typical drinking day, converted to 0, <20, <40, <60, or $\geq 60$ g/day |
| Enough Sleep | Categorical | Perception of getting enough rest from sleep |
| Willingness to change lifestyle | Categorical | Willingness to improve lifestyle habits such as diet, exercise, or smoking |
| Health Interest | Categorical | Interest in improving and maintaining health |

**Supplementary Table 2.** Full baseline characteristics of HHSA and KCMA cohorts.

|  | **HHSA dataset (n = 100,056)** | **KCMA dataset (n = 79,756)** |
| --- | --- | --- |
| **Age, years** | 45 ± 11 | 68 ± 10 |
| **Male sex, n (%)** | 60,957 (60.9%) | 31,260 (39.2%) |
| **Body measurements, mean ±SD** |  |  |
| Height, cm | 166.0 ± 8.6 | 156.8 ± 9.4 |
| Weight, kg | 63.7 ± 13.4 | 56.7 ± 11.0 |
| BMI, kg/m<sup>2</sup> | 23.0 ± 3.9 | 23.0 ± 3.4 |
| Waist, cm | 82.0 ± 10.6 | 83.6 ± 9.7 |
| Systolic blood pressure, mmHg | 122 ± 16 | 129 ± 17 |
| Diastolic blood pressure, mmHg | 74 ± 12 | 75 ± 11 |
| **Blood test, mean ±SD** |  |  |
| Red blood cell count, ×10<sup>6</sup>/μL | 477 ± 47 | 439 ± 46 |
| Hemoglobin, g/dL | 14.3 ± 1.6 | 13.4 ± 1.5 |
| Hematocrit, % | 42.9 ± 4.2 | 40.7 ± 4.0 |
| AST, U/L | 23 ± 15 | 25 ± 21 |
| ALT, U/L | 24 ± 21 | 21 ± 16 |
| GGT, U/L | 38 ± 51 | 36 ± 54 |
| Total Chol, mg/dL | 206± 38 | 205± 35 |
| HDL-C, mg/dL | 62 ± 15 | 59 ± 15 |
| Triglyceride, mg/dL | 114 ± 99 | 125 ± 80 |
| LDL-C, mg/dL | 121 ± 32 | 121 ± 31 |
| Glucose, mg/dL | 96 ± 23 | 103 ± 30 |
| HbA1c, % | 5.6 ± 0.7 | 5.5± 0.7 |
| Creatinine, mg/dL | 0.8 ± 0.2 | 0.7 ± 0.2 |
| eGFR, mL/min/1.73 m<sup>2</sup> | 82 ± 15 | 72 ± 16 |
| **Urine test, n (%)** |  |  |
| Urine Protein |  |  |
| - | 96,839 (96.8%) | 65,168 (81.7%) |
| ± | 2,252 (2.3%) | 9,006 (11.3%) |
| + | 965 (1.0%) | 5,582 (7.0%) |
| Urine Glucose |  |  |
| - | 97,512 (97.5%) | 74,405 (93.3%) |
| ± | 1,667 (1.7%) | 1,615 (2.0%) |
| + | 877 (0.9%) | 3,736 (4.7%) |
| **Electrocardiogram abnormality, n (%)** |  |  |
| Normal | 79,629 (79.6%) | 57,773 (72.4%) |
| Borderline | 12,853 (12.8%) | 14,419 (18.1%) |
| Abnormal | 7,574 (7.5%) | 7,564 (9.5%) |
| **Fundus of eye test, n (%)** |  |  |
| Normal | 77,151 (77.1%) | 77,874 (97.6%) |
| Borderline | 18,049 (18.0%) | 1,001 (1.3%) |
| Abnormal | 4,856 (4.9%) | 881 (1.1%) |
| **Medication, n (%)** |  |  |
| Antihypertensive agents | 9,049 (9.0%) | 31,953 (40.1%) |
| Insulin use | 2,982 (3.0%) | 6,827 (8.6%) |
| Lipid-lowering agents | 3,667 (3.7%) | 19,772 (24.8%) |
| **Self-reported lifestyle, n (%)** |  |  |
| Past Anemia | 5,033 (5.0%) | 11,683 (14.6%) |
| Smoker | 29,491 (29.5%) | 10,423 (13.1%) |
| Weight Gain | 29,242 (29.2%) | 24,161 (30.3%) |
| Regular Exercise | 12,032 (12.0%) | 31,657 (39.7%) |
| Daily Exercise | 21,002 (21.0%) | 42,055 (52.7%) |
| Fast Walking | 22,800 (22.8%) | 34,350 (43.1%) |
| Eating Speed |  |  |
| Slow | 34,366 (34.3%) | 23,117 (29.0%) |
| Normal | 57,260 (57.2%) | 45,596 (57.2%) |
| Fast | 8,430 (8.4%) | 11,043 (13.8%) |
| Late Meals | 24,734 (24.7%) | 14,373 (18.0%) |
| Skip Breakfast | 17,320 (17.3%) | 7,417 (9.3%) |
| Habitual Drinker | 26,736 (26.7%) | 19,386 (24.3%) |
| Daily alcohol intake |  |  |
| None | 51,222 (51.2%) | 29,276 (36.7%) |
| < 20g/day | 20,237 (20.2%) | 34,783 (43.6%) |
| < 40g/day | 18,411 (18.4%) | 10,478 (13.1%) |
| < 60g/day | 8,150 (8.1%) | 3,908 (4.9%) |
| ≥ 60g/day | 2,036 (2.0%) | 1,311 (1.6%) |
| Enough Sleep | 69,125 (69.1%) | 61,919 (77.6%) |
| **Willingness to change lifestyle** |  |  |
| Not willing | 33,306 (33.3%) | 31,794 (39.9%) |
| Somewhat willing | 40,717 (40.7%) | 32,006 (40.1%) |
| Willing | 26,033 (26.0%) | 15,956 (20.0%) |
| **Health Interest, n (%)** | 64,829 (64.8%) | 50,203 (62.9%) |
| **CVD events, n (%)** | 4,113 (4.1%) | 21,179 (26.6%) |
| **Mean follow-up duration, years** | 4.9 ± 7.1 | 7.1 ± 8.0 |

